# *SMPD1* variants do not have a major role in REM sleep behavior disorder

**DOI:** 10.1101/2020.02.15.20023374

**Authors:** Uladzislau Rudakou, Naomi C. Futhey, Lynne Krohn, Jennifer A. Ruskey, Karl Heilbron, Paul Cannon, The 23andMe Research Team, Armaghan Alam, Isabelle Arnulf, Michele T.M. Hu, Jacques Y. Montplaisir, Jean-François Gagnon, Alex Desautels, Yves Dauvilliers, Marco Toffoli, Gian Luigi Gigli, Mariarosaria Valente, Birgit Högl, Ambra Stefani, Evi Holzknecht, Karel Sonka, David Kemlink, Wolfang Oertel, Annette Janzen, Giuseppe Plazzi, Elena Antelmi, Michela Figorilli, Monica Puligheddu, Brit Mollenhauer, Claudia Trenkwalder, Friederike Sixel-Döring, Valérie Cochen De Cock, Christelle Charley Monaca, Anna Heidbreder, Luigi Ferini-Strambi, Femke Dijkstra, Mineke Viaene, Abril Beatriz, Bradley F. Boeve, Ronald B. Postuma, Guy A. Rouleau, Ziv Gan-Or

## Abstract

Mutations in the sphingomyelin phosphodiesterase 1 (*SMPD1*) gene were reported to be associated with Parkinson disease (PD) and dementia with Lewy bodies (DLB). The majority of patients with isolated rapid eye movement sleep behavior disorder (iRBD) develop PD or DLB later in life, suggesting that iRBD is a prodromal phase of these two conditions. In the current study we aimed to evaluate the role of *SMPD1* variants in iRBD. *SMPD1* and its untranslated regions were sequenced using targeted next-generation sequencing in 959 iRBD patients and 1,287 controls from European descent. Logistic regression adjusted for sex and age showed no significant associations with two common variants and iRBD (rs1050239 and rs8164). The frequency of all rare nonsynonymous *SMPD1* variants (minor allele frequency <1%) was found to be twice as high in cases than in controls (1.46% vs. 0.70%, Fisher’s exact test *p=0*.*09*) but there was no statistically significant burden (*p=0*.*64*). Our study reports no statistically significant association of *SMPD1* variants and iRBD. It is hence unlikely that *SMPD1* plays a major role in iRBD.

## 1. Introduction

The sphingomyelin phosphodiesterase 1 (*SMPD1*) gene on chromosome 11 encodes the lysosomal enzyme acid sphingomyelinase, which converts sphingomyelin into ceramide. Homozygous or compound heterozygous mutations in *SMPD1* may cause Niemann-Pick disease type A (NPA) or type B (NPB), lysosomal storage disorders characterized by acid sphingomyelinase (ASM) deficiency and accumulation of sphingomyelin (Schuchman and Desnick, 2017). Sphingomyelin is a major component of both cell membranes and the myelin sheath that surrounds axons in the nervous system (Jana and Pahan, 2010; Quinn, 2014). In recent years, heterozygous *SMPD1* variants have been reported as risk factors for PD (Alcalay et al., 2019; Dagan et al., 2015; Foo et al., 2013; Gan-Or et al., 2013; Mao et al., 2017), and have been suggested to be associated with DLB (Clark et al., 2015).

Rapid-eye-movement (REM) sleep behavior disorder (RBD) is characterized by lack of motor inhibition and dream enactment during REM sleep (St Louis et al., 2017). Accumulating evidence suggests that isolated RBD (iRBD) is a prodromal synucleinopathy, since individuals with isolated RBD are very likely to convert to PD, DLB, or multiple system atrophy (MSA) (Postuma et al., 2019). In order to examine the genetic basis of iRBD and its conversion, recent studies have examined whether PD- or DLB-associated genes are also associated with iRBD and whether they affect its conversion. It was demonstrated that while some genes such as *GBA* (Krohn et al., 2019b), *TMEM175* (Krohn et al., 2019a) and *SNCA* (Krohn et al., 2019c) are associated with iRBD, other PD and DLB genes such as *LRRK2* (Ouled Amar Bencheikh et al., 2018), *APOE* (Gan-Or et al., 2017) and *MAPT* (Li et al., 2018) are not. However, the association between *SMPD1* and iRBD has not been investigated.

Therefore, in the current study we aimed to examine whether rare and common *SMPD1* variants are associated with iRBD. The entire coding regions, the exon-intron boundaries and the 5’ and 3’ untranslated regions of *SMPD1* were sequenced in a large cohort of iRBD patients and controls, followed by different analyses to test for association of specific variants or burden of multiple variants in iRBD.

## 2. Methods

### 2.1. Study Population

A total of 2,246 subjects, composed of 959 unrelated iRBD patients and 1,287 controls, were included in the study. All subjects were of European descent, confirmed by principal component analysis of genotype data that were available for these patients, as we have previously reported (Krohn et al., 2019a). Patients with iRBD had a mean age of 67.9 ± 9.2 (data on age were missing for 7 patients) with 80.7% males (data missing for 31 patients). Controls were all over the age of 40, with a mean age of 59.2 ± 9.3 and 53.7% males. RBD was diagnosed with video-polysomnography according to the International Classification of Sleep Disorders (ICSD), version 2 or 3 (Hogl and Stefani, 2017). All participants signed an informed consent form before entering the study, and the institutional review boards approved the study protocols.

To further investigate one variant in RBD, we examined genome-wide association study (GWAS) summary statistics provided by 23andMe from 1,782 PD cases with probable RBD (PD+pRBD) and 131,250 age- and sex-matched controls. In this cohort, probable RBD was assessed using the RBD1Q questionnaire which has high sensitivity and specificity in PD patients (Postuma et al., 2012). Cases and controls were genotyped on Illumina genotyping platforms, underwent standard GWAS quality control procedures and were imputed and filtered for imputation quality score > 0.80. All individuals included in the analyses provided informed consent and answered surveys online according to the 23andMe human subject protocol, which was reviewed and approved by Ethical & Independent Review Services, a private institutional review board (http://www.eandireview.com). The full protocol is available upon request.

### 2.2. SMPD1 Sequencing

Coding sequence and regulatory regions of *SMPD1* were targeted using molecular inversion probes (MIPs), designed as previously described (O’Roak et al., 2012). MIPs were selected based on their predicted coverage, quality and overlap, and capture was done as previously reported (Ross et al., 2016). Supplementary Table 1 includes all the MIPs used to sequence *SMPD1*, and the full protocol is available upon request. The library was sequenced using Illumina HiSeq 2500 or 4000 platform at the McGill University and Genome Quebec Innovation Centre. Sequence processing was performed by: Burrows-Wheeler Aligner for alignment to the human reference genome (hg19) (Li and Durbin, 2009), the Genome Analysis Toolkit (GATK, v3.8) for post-alignment cleanup and variant calling (McKenna et al., 2010), and ANNOVAR for annotation (Wang et al., 2010). Frequency data on each *SMPD1* variant was extracted from the Genome Aggregation Database (gnomAD) (Lek et al., 2016). Pathogenicity of variants was examined with ClinVar (https://www.ncbi.nlm.nih.gov/clinvar/), especially with respect to Niemann-Pick’s disease. Validations of the rare variants was performed with Sanger sequencing. Supplementary Table 2 details the primers used for validations, and the full protocol is available upon request.

**Table 1.**
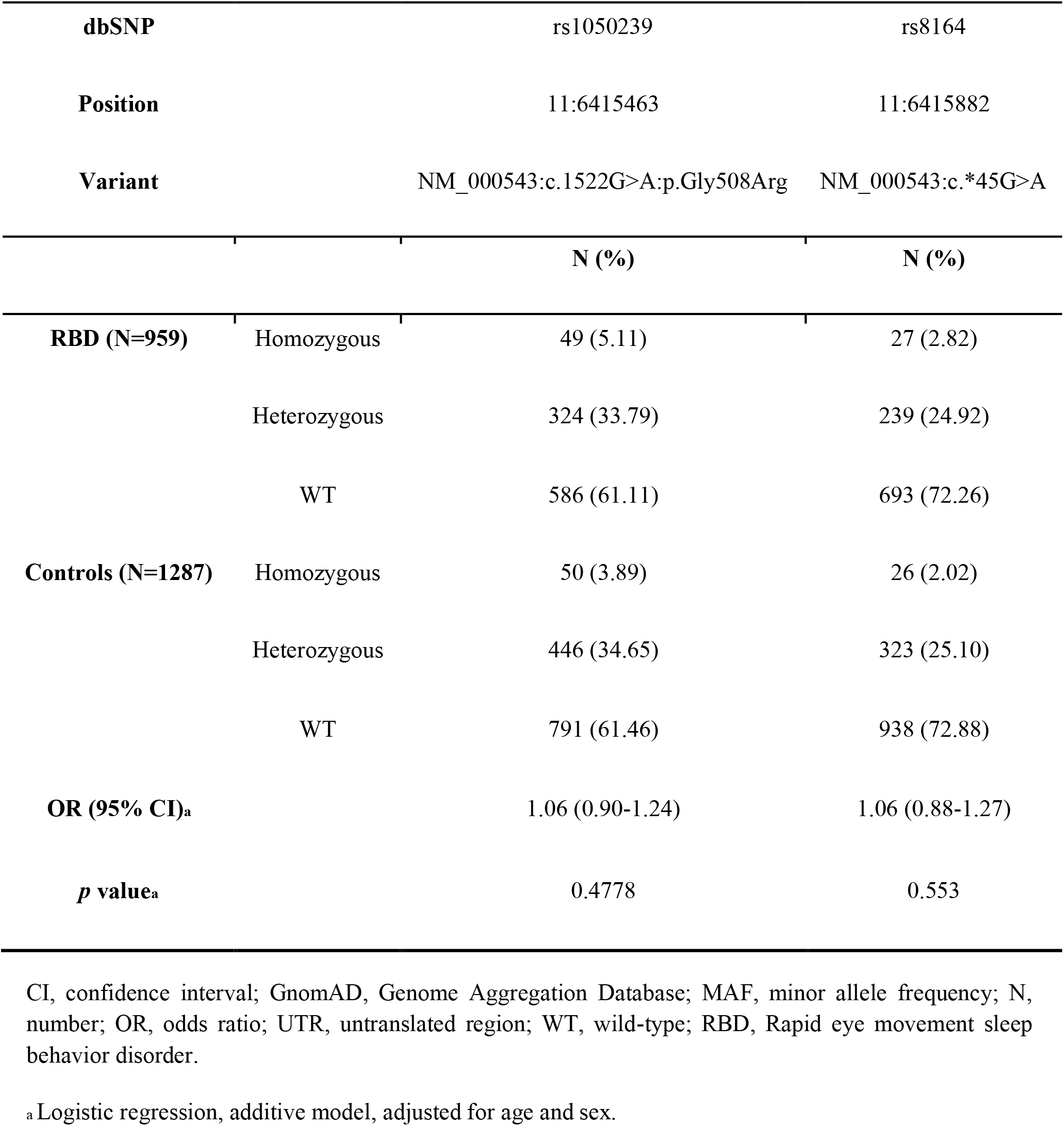
Association of common variants in *SMPD1* with risk for RBD.

| dbSNP |  | rs1050239 | rs8164 |
| --- | --- | --- | --- |
| Position |  | 11:6415463 | 11:6415882 |
| Variant |  | NM_000543:c.1522G>A;p.Gly508Arg | NM_000543:c.*45G>A |
|  |  | N (%) | N (%) |
| **RBD (N=959)** | Homozygous | 49 (5.11) | 27 (2.82) |
|  | Heterozygous | 324 (33.79) | 239 (24.92) |
|  | WT | 586 (61.11) | 693 (72.26) |
| **Controls (N=1287)** | Homozygous | 50 (3.89) | 26 (2.02) |
|  | Heterozygous | 446 (34.65) | 323 (25.10) |
|  | WT | 791 (61.46) | 938 (72.88) |
| **OR (95% CI)<sup>a</sup>** |  | 1.06 (0.90-1.24) | 1.06 (0.88-1.27) |
| ***p* value<sup>a</sup>** |  | 0.4778 | 0.553 |

CI, confidence interval; GnomAD, Genome Aggregation Database; MAF, minor allele frequency; N, number; OR, odds ratio; UTR, untranslated region; WT, wild-type; RBD, Rapid eye movement sleep behavior disorder.
<sup>a</sup> Logistic regression, additive model, adjusted for age and sex.

**Table 2.** Rare nonsynonymous variants and deletions detected in RBD and controls.

| Position | dbSNP | Nucleotide change | Amino acid change | RBD (N=959) | Control (N=1287) | gnomAD MAF (Non-Finnish Europeans) |
| --- | --- | --- | --- | --- | --- | --- |
| 11:6412055 |  | T/C | NM_000534:p.Ile76Thr | 0 | 1 (0.08%) | 0 |
| 11:6413013 |  | C/T | NM_000534:p.Arg240Trp | 1 (0.10%) | 0 | 0 |
| 11:6413367 |  | G/A | NM_000534:p.Glu358Lys | 1 (0.10%) | 0 | 1.80E-05 |
| 11:6413167 | rs1803161 | G/A | NM_000534:p.Arg291His | 2 (0.21%) | 2 (0.16%) | 2.20E-03 |
| 11:6413182 | rs35824453 | G/A | NM_000534:p.Arg296Gln | 1 (0.10%) | 1 (0.08%) | 1.00E-04 |
| 11:6414489 |  | C/T | NM_000534:p.Leu379Phe | 0 | 1 (0.08%) | 6.67E-05 |
| 11:6415245 | rs141641266 | C/T | NM_000534:p.Ala487Val | 8 (0.83%) | 3 (0.23%) | 4.10E-03 |
| 11:6415766 | rs120074118 | TGCC/T | NP_000534.3:p.Arg610del | 1 (0.10%) | 1 (0.08%) | 2.00E-04 |
| **Total** |  |  |  | 14 (1.46%) | 9 (0.70%) |  |

RBD, Rapid eye movement sleep behavior disorder; GnomAD, Genome Aggregation Database; MAF, minor allele frequency; N, number

### 2.3. Quality Control

Quality control (QC) filtration was performed using GATK v3.8 and PLINK software v1.9 (Purcell et al., 2007). Samples with average genotyping rate of less than 90% were excluded. We excluded variants based on the following criteria: coverage quality score (GQ) below 30, minimum depth of coverage which was set to 15x for common variants and 30x for rare variants, genotyping rate of less than 90%, deviation from the Hardy-Weinberg equilibrium set at *p*=0.001, missingness difference between patients and controls set at *p*=0.05 and adjusted by Bonferroni correction.

### 2.4. Statistical Analysis

The association between common *SMPD1* variants and iRBD was tested using a logistic regression using PLINK v1.9, with the disease status (case or control) as a dependent variable, adjusted for age and sex. To analyze rare variants as defined by a minor allele frequency (MAF) of less than 0.01, optimized Sequence Kernel Association Test (SKAT-O, R package) was performed (Lee et al., 2012).

## 3. Results

The average coverage of *SMPD1* was 393x, with 96% of the nucleotides covered at >20x, and 85% covered at >50x. Table 1 details the frequencies of two common variants, rs1050239 and rs8164, in iRBD patients and controls. Both variants were not associated with iRBD (*p=*0.48 and *p=*0.55, respectively, Table 1). Table 2 details rare nonsynonymous and indel variants (no splice or stop variants were identified) with MAF < 0.01 in iRBD patients and controls. The frequency of rare variants in iRBD patients was double than their frequency in controls, but the difference was not statistically significant (1.46% vs. 0.70% respectively, Fisher’s exact test *p=*0.09). There was no statistically significant burden of rare variants in *SMPD1* in iRBD (SKAT-O, *p=*0.64). The difference in frequency of rare variants between RBD patients and control was mainly driven by the p.Ala487Val. This variant was previously nominally associated with PD, albeit non-significant after correction for multiple comparisons (Alcalay et al., 2019), and it was found in 8 (0.83%) of iRBD patients and in 3 (0.23%) of controls (OR=3.6, 95% CI=0.95-13.6, *p*=0.06). However, since the control population in the current study includes controls that were used in the previous study, it is possible that by chance the frequency of this variant in our control population is low. To test this possibility, we examined data from 23andMe, comparing 1,782 PD+pRBD patients and 131,250 controls. The allele frequency of the p.Ala487Val was 0.53% in patients and 0.46% in controls, suggesting a lack of association between this variant and pRBD in PD patients (OR=1.13, 95% CI=0.68-1.88, *p=0*.*64*), and indeed suggesting that the frequency in our control population may be low by chance.

## 4. Discussion

In the current study we fully sequenced *SMPD1* in 959 iRBD patients and 1,287 controls and identified 8 rare *SMPD1* variants (MAF<0.01), and 2 common variants (MAF>0.01). We found no strong evidence for an association of rare or common *SMPD1* variants with iRBD. In a previous large study of PD, the frequencies of *SMPD1* mutations in PD patients and controls of Ashkenazi Jewish (AJ) origin were 1.7% and 0.4%, respectively (Alcalay et al., 2019). With our sample size, we had >80% power to detect such differences between patients and controls at *p*<0.05. However, since our population is less homogeneous than the AJ population, we cannot rule out that rare *SMPD1* variants or variants with small effect size contribute to iRBD, and larger studies will be required in the future to conclusively determine the role of *SMPD1* in iRBD.

Rare *SMPD1* variants were more frequent in cases compared to controls, yet without statistical significance. The increased frequency in patients was driven by the p.Ala487Val variant, which was previously nominally associated with PD, albeit non-significant after correction for multiple comparisons. However, the European control group that was used in the current study included the European controls used in the previous study. Therefore, it is possible that the reduced frequency in the control population is due to chance and drives the apparent association reported in PD and the differences between RBD and controls reported here. To further examine whether p.Ala487Val is associated with RBD, we examined data from 23andMe comparing PD+pRBD patients and controls, which yielded statistically non-significant results. These results may suggest that this variant is not associated with RBD, and may further imply that the previously reported association of this specific variant with PD was also due to chance. However, we cannot rule out a minor role for this variant that requires larger cohorts and statistical power to detect. It is important to note that while this variant is likely not pathogenic in Niemann-Pick disease (Zampieri et al., 2016), it was associated with reduced acid sphingomyelinase activity in dried blood spots from PD patients and controls (Alcalay et al., 2019).

In previous studies, NPA-causing mutations, such as p.L302P (also called p.L304P) and p.fsP330 (also called p.F333Sfs*52 or c.996delC), have consistently been associated with PD (Alcalay et al., 2019; Dagan et al., 2015; Gan-Or et al., 2013). In the current study we did not identify any mutation that causes NPA, but we did identify one mutation, p.Arg610del (previously reported as p.Arg608del or DeltaR608) (Wasserstein and Schuchman, 1993; Zampieri et al., 2016), which causes NPB when inherited in the homozygous or compound heterozygous state with other mutations (Hollak et al., 2012; Jones et al., 2008; Rodriguez-Pascau et al., 2009; Vanier et al., 1993), found in one patient and one control. Therefore, we cannot rule out that rare, thus-far undetected NPA-associated rare variants are associated with iRBD.

Our study has several limitations. Although we analyzed more than 2200 individuals, due to the rarity of NPA-causing mutations we may be underpowered to detect an association of rare *SMPD1* mutations with RBD. Therefore, larger studies will be needed to better define the role of *SMPD1* variants in RBD. Another limitation is that there were differences in age and sex between RBD patients and controls. In the case of rare variants, these differences are not likely to have an effect, and in analyzing common variants we have adjusted for sex and age.

In conclusion, despite the reported association of *SMPD1* variants with PD and DLB, our data does not support a major role for *SMPD1* variants in RBD, yet additional, larger studies are required to conclusively rule out this possibility.

## Data Availability

All data in the manuscript, with the exception of the data from 23andMe, is available upon request.

## Acknowledgements

We thank the patients and control subjects for their participation in this study. This work was financially supported by the Michael J. Fox Foundation, the Canadian Consortium on Neurodegeneration in Aging (CCNA), Parkinson Canada, and the Canada First Research Excellence Fund (CFREF), awarded to McGill University for the Healthy Brains for Healthy Lives (HBHL) program. JFG holds a Canada Research Chair in Cognitive Decline in Pathological Aging. GAR holds a Canada Research Chair in Genetics of the Nervous System and the Wilder Penfield Chair in Neurosciences. WO is Hertie Senior Research Professor, supported by the Charitable Hertie Foundation, Frankfurt/Main, Germany. EAF holds a Canada Research Chair (Tier 1) in Parkinson Disease. ZGO is supported by the Fonds de recherche du Québec - Santé (FRQ-S) Chercheurs-boursiers award, and is a Parkinson’s Disease Canada New Investigator awardee. We thank Daniel Rochefort, Helene Catoire and Vessela Zaharieva for their assistance. Members of the 23andMe Research Team: Michelle Agee, Adam Auton, Robert K. Bell, Katarzyna Bryc, Sarah L. Elson, Pierre Fontanillas, Nicholas A. Furlotte, Barry Hicks, David A. Hinds, Karen E. Huber, Ethan M. Jewett, Yunxuan Jiang, Aaron Kleinman, Keng-Han Lin, Nadia K. Litterman, Matthew H. McIntyre, Kimberly F. McManus, Joanna L. Mountain, Elizabeth S. Noblin, Carrie A.M. Northover, Steven J. Pitts, G. David Poznik, Janie F. Shelton, Suyash Shringarpure, Chao Tian, Joyce Y. Tung, Vladimir Vacic, and Xin Wang

## Conflicts of Interest

KH, PC, and members of the 23andMe Research Team are employees of 23andMe, Inc., and hold stock or stock options in 23andMe. ZGO has received consultancy fees from Idorsia, Denali, Lysosomal Therapeutics Inc. (LTI), Ono Therapeutics, Prevail Therapeutics, Inceptions Sciences (now Ventus) and Deerfield.

